# A high-throughput Epstein-Barr virus nuclear antigen 1 (EBNA1) serology test strip for nasopharyngeal carcinoma risk screening

**DOI:** 10.64898/2026.04.08.26350329

**Authors:** Benjamin E Warner, Japan Patel, Rebecca Satterwhite, Renwei Wang, Jennifer Adams-Haduch, Woon-Puay Koh, Jian-Min Yuan, Kathy H Y Shair

**Affiliations:** Cancer Virology Program, UPMC Hillman Cancer Center, University of Pittsburgh, Pittsburgh, Pennsylvania, USA; Cancer Epidemiology and Prevention Program, UPMC Hillman Cancer Center, University of Pittsburgh, Pittsburgh, Pennsylvania, USA; Department of Epidemiology, School of Public Health, University of Pittsburgh, Pittsburgh, Pennsylvania, USA; Department of Biostatistics, School of Public Health, University of Pittsburgh, Pittsburgh, Pennsylvania, USA; Cancer Risk, Outcomes and Prevention Program, UPMC Hillman Cancer Center, University of Pittsburgh, Pittsburgh, Pennsylvania, USA; Healthy Longevity Translational Research Programme, Yong Loo Lin School of Medicine, National University of Singapore, Singapore; Department of Microbiology and Molecular Genetics, University of Pittsburgh, Pittsburgh, Pennsylvania, USA

## Abstract

**Purpose:** Antibodies to Epstein-Barr virus (EBV) proteins can predict nasopharyngeal carcinoma (NPC) risk. We previously defined a prototype EBNA1 protein panel and multiplex immunoblot assay that distinguishes NPC risk several years pre-diagnosis. Assay throughput and specificity are critical to effectively implement a population-level screening program. Here, we developed a strip test assay – EBNA1 SeroStrip-HT – with an objective to increase throughput and maximize specificity.

**Experimental Design:** EBNA1 full-length (FL) and glycine-alanine repeat deletion mutants (dGAr) were purified from insect and mammalian cells to screen serum IgA/IgG from prospective cohorts in Singapore and Shanghai, China, with known time intervals to NPC diagnosis. Twenty pre-diagnostic sera within 4 years to diagnosis were compared to 96 healthy controls using a nested case-control study design.

**Results:** IgA to mammalian-derived EBNA1 dGAr achieved 85.0% sensitivity and 94.8% specificity (AUC, 0.939) for NPC status. IgA to insect-derived EBNA1 dGAr showed the same sensitivity (85.0%) and similar specificity (93.8%) (AUC, 0.941). IgA to insect-derived EBNA1 FL had a higher 90% sensitivity, but lower 91.7% specificity (AUC, 0.940). Combining EBNA1 FL and dGAr results showed that subjects positive for both proteins had a 243.67 odds ratio for NPC incidence compared to double-negative scores.

**Conclusion:** This study demonstrated the efficacy of EBNA1 SeroStrip-HT for NPC risk assessment and stratification in high- and intermediate-risk populations, yielding high accuracy and a 12-fold increased throughput over the prototype. The insect system was appropriate for large-scale production of purified EBNA1. Larger, geographically diverse cohorts are warranted to confirm these results, especially in low-incidence populations.

## INTRODUCTION

Nasopharyngeal carcinoma (NPC) is a head and neck cancer that is closely associated with Epstein- Barr virus (EBV) infection. More than half of annual NPC diagnoses originate in southern China and Southeast Asia or among first- to third-generation immigrants from those regions (1–5). Early-stage NPC is often subclinical or presents with nonspecific symptoms that delay diagnosis when treatment is favorable (6). Concentrated efforts to develop EBV-based biomarkers are underway that will enable population screening toward improving early diagnosis and longitudinal risk stratification (7).

EBV serology-based screening has shown remarkable accuracy for early-stage NPC diagnosis. In a large screening trial in southern China, an enzyme-linked immunosorbent assay (ELISA) that detects elevated IgA against EBV nuclear antigen 1 (EBNA1) and viral capsid antigen (VCA) p18 identified NPC with 93% sensitivity and 97% specificity (7–9). EBNA1 is a multifunctional protein that is produced in persistent infection because it tethers the viral episome to the host genome (10), whereas VCA p18 is produced only during viral replication (whether permissive or abortive). Measurement of antibodies against these proteins improved early- stage diagnoses by 60% and reduced mortality by 30% among screened participants, including a 44% reduction in those ≥50 years old highlighting the potential health impact for ageing populations (11). However, the positive predictive value (PPV) at 1- and 8-years post-testing was 4.4% and 5.1% respectively, revealing that most screen-positive participants did not develop NPC (7–9). This limitation underscores the need for refined screening assays with high specificity that can evaluate longitudinal NPC risk in screen-positive individuals while preserving cost-effective implementation (7).

New serology assays aim to address barriers to high PPV and longitudinal risk. An ELISA that measures IgA and IgG against BNLF2b is a promising additional EBV target for an NPC screening assay that yields a 44.6% PPV at 6-months but at the cost of a reduced 70.2% sensitivity (12). A high-throughput multiplex bead-based assay that measures IgA and IgG against several EBV lytic and latent proteins can generate a composite antibody signature that captures a broad spectrum of immunity (13–16). This assay has shown similar efficacy in NPC-endemic and non-endemic regions which can enable targeted screening in low- incidence populations that may diverge in EBV-specific immunity (16). The identification of a distinctive pre-diagnostic antibody signature can enrich populations at high risk of NPC for clinical follow-up and stratify risk in screen-positive individuals who remain undiagnosed by endoscopy or MRI (17).

We previously reported that measuring IgA against the cancer-associated EBNA1 sequence variant can predict NPC with 100% sensitivity and 93.7% specificity within 4 years to diagnosis in sera from incident cases from two independent prospective cohorts in Singapore and Shanghai, China (18,19). Our prototype serology assay combined a mammalian expression library of 86 EBV proteins with a denaturing multiplex immunoblot to measure serum IgA and IgG (18). Compared to other antibody detection platforms, visual inspection of protein bands provides rigorous scoring criteria but is limited by throughput. In a follow-up study, an expanded EBNA1 library was developed to map epitopes that are critical for predicting NPC risk which demonstrated that polymorphic residues within certain epitopes affected assay sensitivity and specificity (19). Genetic studies have classified EBV sequence heterogeneity by geography and associated disease (20,21), such as the high, intermediate, and low-risk haplotypes in EBV *BALF2* that have been estimated to contribute to 80% of risk in NPC-endemic regions (22–26). Measuring antibodies against the appropriate cancer-associated sequence and epitope could be critical for optimizing risk screening across diverse populations (19). Indeed, we found that IgA binding profiles against EBNA1 were defined by the glycine-alanine repeats (GAr) and C-terminus which categorized pre-diagnostic sera into high-, intermediate-, and low-risk profiles according to the number of years to diagnosis (19). These data suggest that a curated EBNA1 panel in a high-throughput platform could improve NPC early detection and risk assessment in endemic regions.

In this study, our objective was to refine the EBNA1 protein panel and denaturing multiplex immunoblot assay into a scalable test strip while preserving test specificity. The prototype assay measured serum antibodies against whole-cell lysates from mammalian cells transfected with an NPC-associated EBNA1 sequence; the lysates were separated by SDS-PAGE and detected by western blot, a format modeled on the accuracy of original HIV confirmatory assays (27). Here, a slot blot approach was adopted and refined to use three purified EBNA1 protein analytes produced in insect or mammalian cells. Full-length EBNA1 from a baculovirus-insect cell expression system (insEBNA1 FL) and EBNA1 mutants deleted for the glycine-alanine repeat in both insect and mammalian systems (insEBNA1 dGAr and mamEBNA1 dGAr, respectively) were spotted onto nitrocellulose strips and incubated with pre-diagnostic sera for multiplexed fluorescent detection of IgA and IgG. Participants from two independent prospective cohorts in Singapore and Shanghai, China were surveyed using in a nested case-control study design to evaluate the efficacy of the new EBNA1 SeroStrip-HT for NPC risk prediction and stratification.

## MATERIALS AND METHODS

### Study Population and Design

The Singapore Chinese Healthy Study (SCHS) and Shanghai Cohort Study (SCS) have been previously described (18,19). Briefly, the SCHS is a residential cohort of 63,257 Chinese men and women from Singapore, aged 45 to 74 years at enrollment (1993–1998) (28). The SCS is a residential cohort of 18,244 men from Shanghai, China, aged 45 to 64 years at enrollment (1986–1989) (29). Pre-diagnostic sera from 13 SCHS and 7 SCS participants who later developed NPC within 4 years of sample collection were available for this study. The NPC cases evaluated were presumed to be EBV-associated due to the geography and ethnicity of the participants. A nested case-control study design with 5 healthy control subjects to one case was employed. One of the five controls matched to the index cases in our previous studies were retained (18,19). For each case, we successfully selected additional 4 random controls among the cohort participants who were free of cancer and alive at the index date with the same matching criteria on all except for one case. The matching criteria to the SCHS index case were sex, dialect group (Hokkien and Cantonese), age at enrollment (±3 years), and date of biospecimen collection (±6 months). The matching criteria to the SCS index case were age at enrollment (±2 years), date of blood draw (±1 month), and the same neighborhood of residence at study enrollment. In total, there were pre-diagnostic sera <4 years from blood collection to diagnosis of NPC and 96 healthy control subjects available for this study. All participant samples in this study were collected once, and none were resampled longitudinally.

### EBNA1 Insect and Mammalian Expression Vectors

EBNA1 constructs were expressed as a single open reading frame with a Tobacco Etch Virus (TEV) protease cleavable N-terminal 6xHis-tag followed by a 3xFLAG-tag fused to EBNA1 Akata (GenBank KC207183.1) as full-length protein containing a.a. 1-597 or deletion mutant missing the glycine-alanine repeat (a.a. 90-283). For the baculovirus-insect cell expression system, pFastBac1 expression vectors were developed for insEBNA1 FL and insEBNA1 dGAr (GenScript Biotech, New Jersey). For the mammalian expression system, p3xFLAG-CMV-7m was described previously and used to express mamEBNA1 dGAr (18).

### Cell Line and Purification of Lysates

Spodoptera frugiperda (Sf9) suspension cells were grown in serum-free SF900II medium and incubated at 27°C, 5% CO_2_, in a vented flask on a rotating platform at 125 rpm. Whole-cell lysates for EBNA1 protein purification were prepared as follows: 1×10^6^ Sf9 cells/mL in log growth phase were infected with recombinant baculovirus containing insEBNA1 FL or insEBNA1 dGAr at 1 plaque forming unit/cell for 72 h. The culture was pelleted for 10 min at 4500 rpm and chemically lysed in ice-cold buffer (50 mM Tris-HCl, 300 mM NaCl, 5% glycerol, 1% NP-40, 0.476mg/L Benzonase nuclease, pH 8.0) supplemented with cOmplete Protease Inhibitor Cocktail EDTA-free (Roche). Whole-cell lysates were equilibrated (50 mM Tris-HCl, 300 mM NaCl, 5 % Glycerol, pH 8.0) and applied to a 1 mL Ni-NTA column (GenScript L00250). Columns were washed twice, first in wash buffer 1 (50 mM Tris-HCl, 500 mM NaCl, 5 % Glycerol, 0.5% Triton X-100, 0.5% Triton X-114, pH 8.0) followed by wash buffer 2 (50 mM Tris-HCl, 300 mM NaCl, 5 % Glycerol, pH 8.0). The column was eluted (50 mM Tris-HCl, 300 mM NaCl, 5 % Glycerol, variable imidazole, pH 8.0) in a series of increasing imidazole concentrations of 20, 50, 250, 500 mM. The 250 mM imidazole elution fraction was dialyzed (JKHD MD25-14) and the His-tag was removed by TEV protease (GenScript Z03030-1K) cleavage at a site between the His- and FLAG-tag. The sample was applied to a new 1mL Ni-NTA column to capture the cleaved fragment and protease, and the flow-through was stored at -80°C. The total yield from 500 mL original culture was 0.7 mg insEBNA1 FL and 1.8 mg insEBNA1 dGAr at an estimated 90% purity by SDS-PAGE.

Expi293F suspension cells (ThermoFisher Scientific) were grown in Expi293 Expression Medium (ThermoFisher Scientific) and incubated according to manufacturer protocol at 37°C, 8% CO_2_, in a vented flask on a rotating platform at 120 rpm. Thawed cells were maintained at 95% viability for a maximum of 30 passages. Whole cell lysates for EBNA1 protein purification were prepared by polyethylenimine transfection. Briefly, 3×10^6^ cells/mL at 95% viability were transfected with 1 µg/mL EBNA1 expression vector and 9 µg/mL polyethylenimine 25 kDa (Kyfora Bio) and incubated for 72 h. Cells were pelleted for 20 min 1000 *xg* at 4°C and lysed in ice-cold lysis buffer (50 mM Tris-HCl, 300 mM NaCl, 5% glycerol, 1% NP-40, pH 8.0) freshly supplemented with 0.1 µL/mL nuclease (Pierce, Sigma-Aldrich), 1X HALT protease and 1X HALT phosphatase inhibitor cocktails (Sigma-Aldrich), 0.1 M phenylmethylsulfonyl fluoride, and 0.2 M activated sodium vanadate. The whole-cell lysates were rocked for 30 min at 4°C followed by clarification for 30 min 20,000 *xg* at 4°C. EBNA1 was purified from the lysate using a Pierce Ni-NTA Magnetic Agarose Beads (ThermoFisher Scientific) according to manufacturer protocol with slight modification to the elution procedure. Briefly, the 4 mL bead slurry was separated into two fractions in 15 mL tubes and equilibrated twice in 8 mL buffer (50 mM sodium phosphate, 300 mM NaCl, 10 mM imidazole, 0.05% Tween-20, pH 8.0) and separated on a magnetic stand. The whole-cell lysate was diluted 1:1 in equilibration buffer and each bead fraction was incubated with 14 mL diluted lysate overnight at 4°C on an end-over-end rotator. The beads were separated on a magnetic stand, and each fraction was washed twice in 14 mL buffer (50 mM sodium phosphate, 300 mM NaCl, 15 mM imidazole, 0.05% Tween-30, pH 8.0) before eluting. Elution (50 mM sodium phosphate, 300 mM NaCl, variable imidazole) was carried out in a series of two 7 mL fractions each in increasing imidazole concentrations at 20 mM, 50 mM, 250 mM, and 500 mM. The two 7 mL x 250 mM imidazole fractions were combined and stored at -80°C. The total yield from 300 mL original culture was 0.75 mg at 85% purity by SDS-PAGE.

### EBNA1 SeroStrip-HT Preparation and Serum Survey

Purified EBNA1 protein was applied directly to a nitrocellulose membrane using the BioDot-SF vacuum manifold apparatus (Bio-Rad) according to manufacturer protocol. Briefly, mamEBNA1 dGAr and insEBNA1 dGAr were diluted in PBS to 0.25 ng/ µL supplemented with 2.5% β-mercaptoethanol, boiled for 5 min, and stored at -80°C until loading. The insEBNA1 FL protein was prepared the same way at 0.315 ng/µL to ensure each analyte was loaded at equimolar concentration. Nitrocellulose membranes and filter paper were saturated in 1X tris-buffered saline (TBS) for 10 min. The filter paper was loaded onto the apparatus followed by the membrane and the sample template was screwed into place under vacuum flow. The membrane was washed with 200 µL 1X TBS per well and 200 µL of purified EBNA1 protein applied to specified wells (load total equivalent to 50 ng mamEBNA1 dGAr, 50 ng insEBNA1dGAr, and 63 ng insEBNA1 FL). The wells were washed twice with 200 µL 1X TBS per well and the apparatus was disassembled under vacuum flow. The nitrocellulose membrane was blocked for 1 h in 5% nonfat dry milk in 1xTBS and cut into vertical strips. Separate standard curve blots were produced for purified human IgA and IgG (ChromPure, Jackson IR). Briefly, 24 concentrations of IgA (range 0.780-146.25 ng) or IgG (range 0.950-178.13 ng) were diluted in 200 µL 1X PBS and loaded onto a nitrocellulose membrane following the same procedure noting that the linear range of detection was 2.0–429.0 ng for the IgA detection channel and 7.1–332.5 ng for the IgG detection channel.

After blocking, the test strips were immediately transferred to multi-well trays and incubated overnight at 4°C in a 1:500 dilution of participant serum with 1 µg/mL anti-FLAG M2 clone (AB_259529, Sigma-Aldrich) in blocking buffer. Following overnight incubation, the strips were washed in 1x TBS (3x 5 m) and incubated for 1 h at room temperature in the dark with fluorescent tagged secondary antibodies diluted in blocking buffer [3.75 mg/mL Cy3-AffiniPure goat anti-hIgA_H_ (AB_2337721, Jackson IR); 0.03 mg/mL AlexaFluor 680-AffiniPure goat anti-hIgG_H_ (AB_2889013, Jackson IR); 0.1 mg/mL IR800CW goat anti-mIgG_H+L_ (AB_621842, LI-COR)]. The strips were washed in 1x TBS (3x 5 m) and immediately imaged on a ChemiDoc MP (Bio-Rad) scanner. The levels of detection in each channel were quantified by densitometry using ImageLab 6 software (Bio-Rad). The standard curve blots were treated identically but without incubation of serum. Absolute quantification was calculated according to IgA and IgG standard curve blots and were within the linear range of detection. Personnel running and scoring the test strips were masked to serum identity and case control status.

### EBNA1 Genotyping by Amplicon Sequencing

Cell-free DNA was extracted from 200 µL (pre-dx NPC 1.80y) or 400 µL (pre-dx NPC 0.06y and 2.07y) pre-diagnostic sera using the GeneJET Genomic DNA Purification Kit (Thermo Fisher K0722) with elution volumes of 100 or 200 µL Milli-Q water, respectively. The extractions yielded 20-40 ng of DNA to PCR amplify and genotype the EBNA1 gene. Amplicons were designed to span the known polymorphic site at residue 487 (GenBank ref. V01555.2, Forward primer 5’-AGGCCATTTTTCCACCCTGT-3’, Reverse primer 5’- GTCCAGGGGCCATTCCAAA-3’, yielding a 393 bp amplicon).

A two-step PCR protocol was used to generate sufficient material for sequencing. All PCR reactions used Q5 High-Fidelity 2x MasterMix (NEB M0492L) with 500 nM primers and 68°C annealing temperature. PCR step 1 amplified 1 µL of purified DNA as template for 25 cycles. PCR Step 2 amplified 5 µL step 1 PCR product for 40 cycles. Amplicon size was confirmed by gel electrophoresis and bands were gel purified by the GeneJET Gel Extraction Kit (Thermo Fisher K0692). DNA was quantified by a Qubit 3 Fluorometer (Thermo Fisher Q3316) and samples were normalized to 20 ng/µL prior to amplicon sequencing by the Amplicon-EZ sequencing service (Genewiz, Azenta Life Sciences). Sequencing yielded >10^5^ paired-end 250 bp reads per sample, and run quality was confirmed with FastQC v0.12.0 (Andrews 2010). Variant calling was performed using breseq v0.38.1 (Deatherage and Barrick 2014) with the -p flag to indicate polymorphism mode and the -t flag to indicate targeted sequencing mode.

### Statistical Analysis

Replicate variability was assessed by calculating the coefficient of variation (CV) from the absolute values of duplicate measurements of analytes; mamEBNA1 dGAr (median 3.37, range 0.04-27.43), insEBNA1 dGAr (median 3.23, range 0.03-15.16), and insEBNA1 FL (median 4.68, range 0.01-32.49). A total of 12 duplicate measurements (3.4%) had a CV >20% (1 pre-dx sera from mamEBNA1 dGAr, 4 pre-dx sera from insEBNA1 FL, and 7 healthy controls from insEBNA1 FL). Excluding these samples did not change our conclusions and all data were included in subsequent analyses. Receiver operating characteristic (ROC) curves were generated in R (v4.4.1) using the pROC package (v1.19.0.1) for each EBNA1 construct. Optimal cutoffs were determined by maximizing Youden’s index yielding IgA cutoffs (abs. value): 1:5 matching ratio analysis, mamEBNA1 dGAr IgA; 10.47, insEBNA1 dGAr IgA; 9.29, insEBNA1 FL; 51.96, and IgA cutoffs (abs. value): 1:1 matching-ratio, mamEBNA1 dGAr IgA; 9.60, insEBNA1 dGAr IgA; 8.42, insEBNA1 FL; 50.67 to calculate the sensitivity, specificity, area under the curve (AUC), odds ratio (OR), and 95% confidence interval (95% CI). A Fisher exact test was used to examine the difference in the proportion of serum samples detecting each construct between cases and controls. A simple linear regression was used to determine the coefficients of determination (R^2^) and 95% confidence bands.

EBNA1 risk profiles were determined by comparing IgA detection values of multiple analytes. IgA to insEBNA1 FL and mamEBNA1 dGAr were first independently scored as test-positive or -negative based on the analyte cutoff, then each participant was categorized four profiles: Profile 1 showed IgA absolute values below cutoff for both mamEBNA1 dGAr (cutoff; 10.47) and insEBNA1 FL (cutoff; 51.96) which indicated low antibody detection independent of EBNA1 epitope availability. Profile 2a showed IgA above cutoff for insEBNA1 FL only indicating that high detection is dependent on GAr epitopes. Profile 2b showed IgA above cutoff for mamEBNA1 dGAr only indicating that high detection is independent of GAr epitopes. Profile 3 showed IgA detection above cutoff for both insEBNA1 FL and mamEBNA1dGAr. The OR and 95% confidence interval (CI) were calculated for each profile using Profile 3 as reference (OR=1). Profile 2a and 2b were combined into a single profile for OR calculations. Statistical analyses were performed in R and Prism 10 software. A *P* value of <0.05 was considered statistically significant.

### Study Approval

All study participants provided written informed consent (28,29). The study was conducted in accordance with the ethical guidelines of the Declaration of Helsinki. Approval was granted by the Institutional Review Board (IRB) at the University of Pittsburgh under STUDY19080227 “The Shanghai and Singapore Cohort Studies”, and IRB STUDY24030062 “EBV and NPC risk” (exempt, not human subjects research). The cohorts have been approved by the Institutional Review Boards of the National University of Singapore (for the SCHS) and the Shanghai Cancer Institute (for the SCS) and are compliant with the United States Department of Health and Human Services.

### Data Availability

EBNA1 SeroStrip-HT data and the EBNA1 ORF library can be obtained upon reasonable request. Requests may be subject to institutional approval associated with US patent application serial no. PCT/US23/66374 (pub. no. US 20250389720 A1). In compliance with the National Institutes of Health (NIH) policy for Data Management & Sharing Plan, raw and processed data have been archived in a generalist repository, the Open Science Framework, and are available for download (https://doi.org/10.17605/OSF.IO/34976).

## RESULTS

A total of 20 participants who later developed NPC within 4 years of sample collection and 96 healthy control subjects were selected from the SCHS and SCS (see baseline characteristics in **Supplementary Table 1**). Participant sera were surveyed for IgA and IgG against denatured EBNA1 proteins using the newly developed nitrocellulose test strip (EBNA1 SeroStrip-HT) (**Figure 1** and **Supplementary Figure 1**). Each strip contained three analytes and one background buffer control in duplicate, including mamEBNA1 dGAr, insEBNA1 dGAr, insEBNA1 FL, and phosphate-buffered saline (PBS). EBNA1 purification from mammalian whole cell lysates was adapted from prior studies (18,19). Purification from whole-cell lysates of baculovirus-infected insect cells was guided by prior studies that established the method to produce abundant EBNA1 protein for immunologic analyses (30,31). Each analyte was tagged with 3xFLAG on the N-terminus and the strips were co-incubated with anti-FLAG monoclonal antibodies to control for successful loading (**Supplementary Figure 1**). The buffer control verified that the staining procedure did not produce unexpected fluorescent background. The signal intensity for EBNA1 IgA and IgG was quantified by densitometry. To convert raw fluorescent intensity into absolute quantification, standard curves were prepared with known quantities of purified human IgA and IgG monomers and stained with the same fluorescent secondary antibodies (**Supplementary Figure 2**).

**Figure 1.**
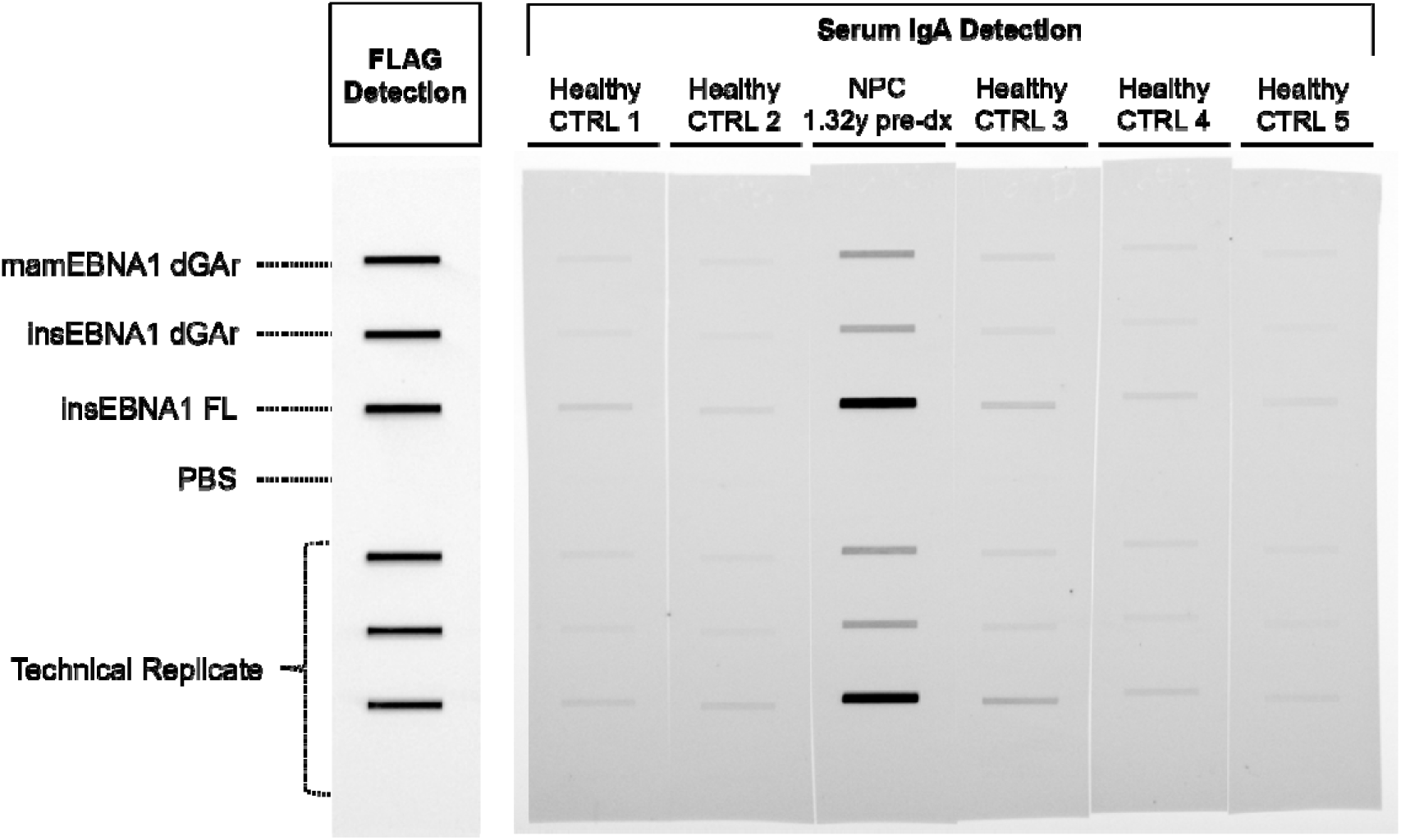
EBNA1 SeroStrip-HT schematic and example of IgA detection. One matched set that included one pre-diagnostic (pre-dx) NPC serum and 5 matched healthy control sera were urveyed simultaneously for IgA, IgG, and FLAG detection of multiple EBNA1 analytes; mammalian-derived EBNA1 deleted for the glycine alanine repeat (mamEBNA1 dGAr) or insect-derived (insEBNA1 dGAr), insect-derived full-length EBNA1 (insEBNA1 FL). The IgA channel of the pre-diagnostic serum 1.32 years to diagnosis is displayed adjacent to matched healthy control sera. See Supplementary Figure 1 for IgG channel and complete FLAG channel.

For each analyte, the absolute quantification of EBNA1 IgA and IgG detection were arranged into heatmaps and ordered by years to diagnosis (**Figure 2**). Consistent with previous findings, EBNA1 IgG was detected uniformly across all participants reflecting the ubiquity of EBV seroconversion in our cohorts (18,19). EBNA1 IgA detection better distinguished participants who developed NPC from cancer-free controls. Removal of the GAr residues reduced overall IgA and IgG detection intensity (**Supplementary Figure 3**). We have previously shown the GAr is a source of strong antibody reactivity in most pre-diagnostic sera and a minority of healthy control subjects that contributed to a reduced specificity for predicting NPC (19). IgA technical replicates were compared to validate test reproducibility and high correlation was observed for each analyte: mamEBNA1 dGAr (R^2^, 0.9921), insEBNA1 dGAr (R^2^, 0.9955), and insEBNA1 FL (R^2^, 0.9895) (**Supplementary Figure 4**). Greater replicate variability was observed at lower IgA detection intensity and 3.4% of replicates yielded a coefficient of variation (CV) >20% including 1 NPC incident case from mamEBNA1 dGAr and 4 NPC and 7 healthy controls from insEBNA1 FL. Excluding these measurements did not alter our conclusions and all case-control pairs were included in the statistical analysis. We also compared the IgA detection of mamEBNA1 dGAr and insEBNA1 dGAr which differed only by the host expression system. Strong correlation mamEBNA1 dGAr and insEBNA1 dGAr was observed among incident NPC cases (R^2^, 0.9934) and healthy controls (R^2^, 0.9848) with greater variation observed at lower detection intensities (**Supplementary Figure 5**).

**Figure 2.**
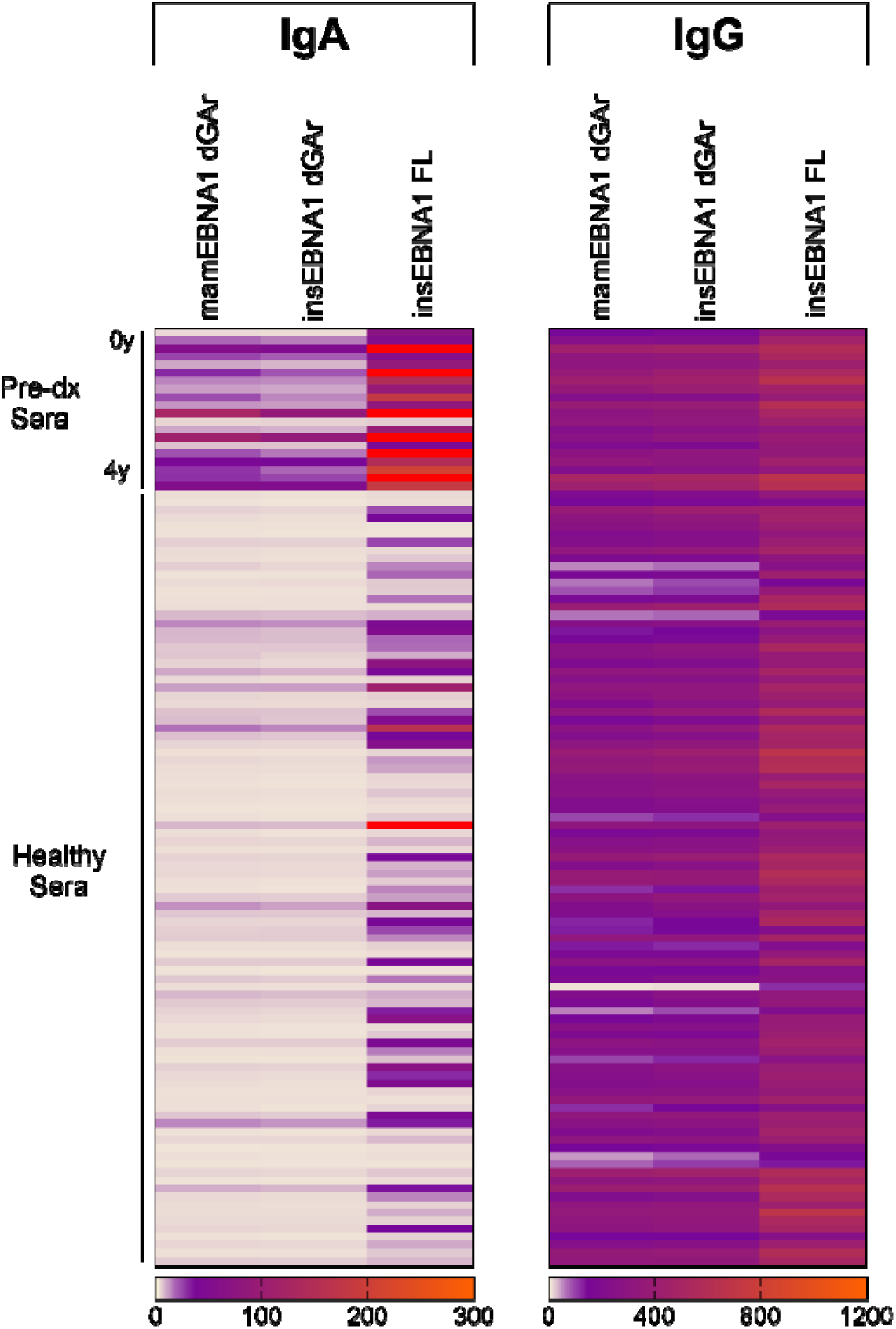
Detection of EBNA1 IgA and IgG for all pre-diagnostic and healthy sera. Pre-diagnostic (pre-dx) sera from 20 incident NPC cases and 96 healthy control subjects were surveyed for detection of IgA (left) and IgG (right) against EBNA1 analytes from EBNA1 SeroStrip-HT. Pre-dx sera were ordered by the number of years to diagnosis. Healthy sera were grouped and ordered according to the matched pre-dx sera. Relative scale for: IgA; tan (0), purple (30), orange (300), red (above scale), IgG; tan (0), purple (120), orange (1200).

Receiver operating characteristic (ROC) curves were assembled to calculate maximum Youden’s index and establish unbiased cutoffs for each analyte (**Supplementary Figure 6**). IgA detection of insEBNA1 FL exceeded the cutoff in 90.0% of pre-diagnostic sera and 8.3% of healthy control subjects. When the GAr sequence was absent, IgA detection of mamEBNA1 dGAr was above cutoff in 85.0% of pre-diagnostic sera and 5.2% of healthy controls. Consequently, mamEBNA1 dGAr yielded the highest overall specificity of 94.8% with a sensitivity of 85.0% (AUC 0.939) (**Table 1**). IgA against insEBNA1dGAr was similar with 93.8% specificity and 85.0% sensitivity (AUC, 0.941), while insEBNA1 FL yielded 91.7% specificity and 90.0% sensitivity (AUC, 0.940). Three NPC cases sampled 0.06, 1.80, and 2.07 years pre-diagnosis fell below the IgA detection cutoff for the EBNA1 dGAr analytes. Inspection of the scans revealed no indication of technical anomaly. In our prior survey using the prototype assay with mamEBNA1 dGAr as a whole-cell lysate, IgA levels for these same sera clustered very close to the optimized cutoff, so the false-negative detection here is not surprising (19). To facilitate the direct comparison of the prototype assay to EBNA1 SeroStrip-HT, ROC curves were assembled for mamEBNA1 dGAr using the original 1:1 case-control matching scheme previously published in the prototype assay (**Supplementary Figure 7**). Setting optimal cutoffs as determined by Youden’s index (**Table 2**), EBNA1 SeroStrip-HT reached 100% specificity for the dGAr analytes (mamEBNA1 dGAr and insEBNA1 dGAr) and 85% sensitivity, while the prototype assay reached maximal 100% specificity and 90% sensitivity with the FL analyte (mamEBNA1 FL). IgA detection for three cases with 0.06, 1.80, and 2.07 years pre-diagnosis were just above cutoff in the prototype assay and the discrepancy of these three samples scoring false-negative in the present study is within expected variance. Genotypes of EBNA1 were successfully determined for two of the three false-negative samples based on their circulating cell free DNA (cfDNA) and determined to be EBNA1 Akata (487V genotype) by majority in the plasma or serum of the indexed case (**Supplementary Table 2**). The dominant EBNA1 strain variant detected in the blood of the false-negative samples matched the EBNA1 variant in the analyte. Thus, the false-negative samples are likely attributed to the limit of detection of the assay and not due to mismatched epitopes.

**Table 1.**
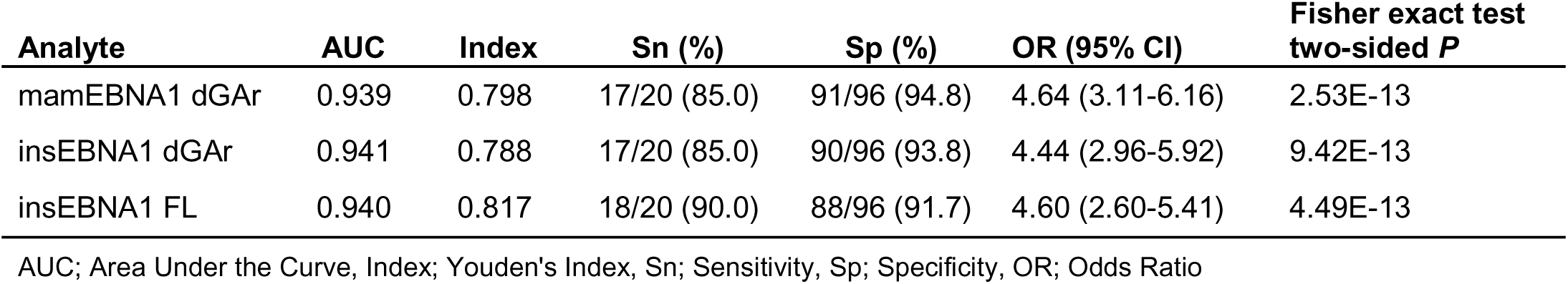
Sensitivity and specificity of EBNA1 IgA by EBNA1 SeroStrip-HT in a 1:5 case-control matching ratio.

| Analyte | AUC | Index | Sn (%) | Sp (%) | OR (95% CI) | Fisher exact test<br>two-sided *P* |
| --- | --- | --- | --- | --- | --- | --- |
| mamEBNA1 dGAr | 0.939 | 0.798 | 17/20 (85.0) | 91/96 (94.8) | 4.64 (3.11-6.16) | 2.53E-13 |
| insEBNA1 dGAr | 0.941 | 0.788 | 17/20 (85.0) | 90/96 (93.8) | 4.44 (2.96-5.92) | 9.42E-13 |
| insEBNA1 FL | 0.940 | 0.817 | 18/20 (90.0) | 88/96 (91.7) | 4.60 (2.60-5.41) | 4.49E-13 |

AUC; Area Under the Curve, Index; Youden's Index, Sn; Sensitivity, Sp; Specificity, OR; Odds Ratio

**Table 2.** Comparison of EBNA1 IgA detection from EBNA1 SeroStrip-HT versus the prototype multiplex immunoblot assay using the 1:1 case-control matching ratio reported in Warner *et al.* 2024 (19).

| Assay | Analyte | AUC | Index | Sn (%) | Sp (%) | OR (95% CI) | Fisher exact test two-sided *P* |
| --- | --- | --- | --- | --- | --- | --- | --- |
| EBNA1 SeroStrip-HT | mamEBNA1 dGAR<sup>a</sup> | 0.963 | 0.850 | 17/20 (85.0) | 20/20 (100.0) | 5.32 (2.29-8.35) | 2.57E-08 |
|  | insEBNA1 dGAR | 0.965 | 0.850 | 17/20 (85.0) | 20/20 (100.0) | 5.32 (2.29-8.35) | 2.57E-08 |
|  | insEBNA1 FL | 0.953 | 0.850 | 18/20 (90.0) | 18/20 (90.0) | 5.14 (2.66-7.63) | 5.82E-08 |
| EBNA1 IgA denaturing multiplex immunoblot<sup>19</sup> | mamEBNA1 dGAR<sup>b</sup> | 0.995 | 0.950 | 20/20 (100.0) | 19/20 (95.0) | 6.28 (3.02-9.54) | 3.05E-10 |
|  | mamEBNA1 FL | 0.955 | 0.900 | 18/20 (90.0) | 20/20 (100.0) | 5.72 (2.61-8.82) | 3.35E-09 |

AUC; area under the curve, Index; Youden's Index, Sn; sensitivity, Sp; specificity, OR; Odds Ratio <sup>a</sup>Purified EBNA1 dGAR from mammalian cells, <sup>b</sup>Whole-cell lysate from mammalian cells containing EBNA1 dGAR

Lastly, for each analyte, EBNA1 IgA and IgG detection was evaluated as a series of sequential or simultaneous testing models to determine if any combination of measurements could improve test accuracy over IgA against mamEBNA1 dGAr. No sequential test model (two-step) outperformed the sensitivity and specificity of mamEBNA1 dGAr alone, but surprisingly, IgG measurements were among the top performing candidates for the second step measurement. For example, IgA against mamEBNA1 dGAr followed by IgG detection against insEBNA1 FL or mamEBNA1 dGAr each yielded 75% sensitivity and 100% specificity suggesting IgG to EBNA1 can add value to NPC prediction despite the ubiquity of EBNA1 IgG systemic immunity in the study population. Simultaneous testing models marginally improved specificity when combining IgA against insEBNA1 FL with mamEBNA1 dGAr (96.9% specificity at 85% sensitivity) as compared to mamEBNA1 dGAr alone (94.8% at 85% sensitivity). More importantly, this test model efficiently categorized participants into NPC high-, intermediate-, and low-risk profiles based on reactivity to the GAr and C-terminus (**Table 3** and **Supplementary Figure 8**), which we previously reported for 79 pre-diagnostic sera up to 23.92 years to diagnosis using the prototype assay (19). The high-risk profile is defined by IgA detection of EBNA1 of both insEBNA1 FL and mamEBNA1 dGAr which confirmed that selective reactivity to non-repetitive sequences outside of the GAr are important for NPC risk assessment. The majority of sera within 4 years to diagnosis (n=17/20) profiled as high-risk in the EBNA1 SeroStrip-HT IgA assay compared to three healthy controls (n=3/96). In contrast, the intermediate-risk profile is defined by IgA reactivity to either the GAr sequence or unique non-repetitive sequences which distinguished a minority of pre-diagnostic sera (n=1/20) and healthy controls (n=7/96). The low-risk profile that shows weak detection across all EBNA1 analytes was characteristic of most healthy controls (n=86/96) compared to few NPCs (n=2/20). We calculated the follow-up time for healthy controls subjects which indicated that the controls who profiled high- or intermediate-risk were unlikely to have undiagnosed NPC after an average of 14-17 years of follow-up (**Table 3**).

**Table 3.**
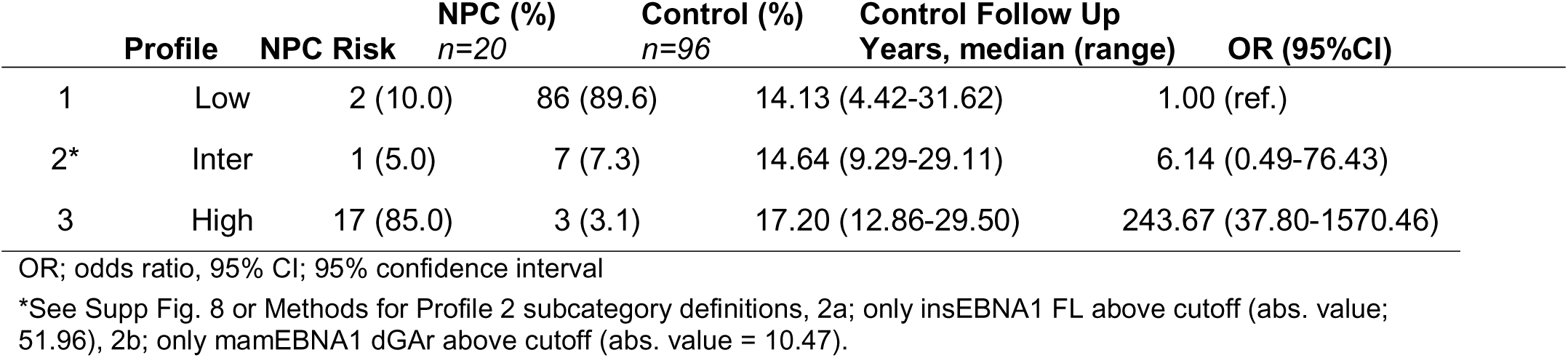
Profile of NPC risk based on simultaneous detection of IgA against insEBNA1 FL and mamEBNA1 dGAr.

## DISCUSSION

A one-time EBV serology screening test is cost effective in NPC-endemic populations and is recommended for ethnic groups at high- or intermediate-risk of developing NPC (7,32). Converting EBNA1 SeroStrip-HT into a standardized test could serve as a reflex test for existing NPC early detection programs that would prioritize test-positive individuals with unconfirmed cancer status for continued clinical follow up who remain at elevated risk for NPC. Alternatively, it may be implemented in a new risk assessment program to prioritize who should undergo routine early detection screening. EBNA1 SeroStrip-HT retained many hallmarks of our prototype assay including consistent detection of EBNA1 IgG across cases and controls that indicated EBV seropositivity, and the ability to profile NPC risk by time to diagnosis. Elevated levels of EBNA1 IgA with distinct binding profiles to the GAr and/or the non-repetitive C-terminal sequences highlights a defining feature of EBNA1 SeroStrip-HT over other serology assays that do not leverage differential epitope binding to infer risk.

Given that NPC incidence ranges between 25-30 per 100,000 persons in NPC-endemic areas to 0.5 per 100,000 in non-endemic areas, optimizing test specificity remains a priority (7). The mamEBNA1 dGAr analyte achieved a maximum 94.8% specificity for discriminating pre-diagnostic sera and insEBNA1 dGAr yielded a comparable 93.8% specificity. Due to the repetitive immunodominant epitopes in the GAr, insEBNA1 FL showed the strongest overall IgA signal but the lowest specificity of 91.7%. Despite promising avenues for NPC screening that measure antibody diversity, an EBV and host risk haplotype composite score, and/or unique characteristics of plasma cfDNA, there remains an unmet need to risk stratify in test-positive individuals who remain tumor-negative after endoscopy or MRI (8,22,24,33). Efforts have indeed been attempted in a prospective NPC-endemic cohort study using EBV cfDNA fragmentomics profiling to distinguish individuals who tested positive for elevated EBV cfDNA and were diagnosed with NPC within 4 years from participants who remained cancer-free (relative risk 87.1) (34). In our 1:5 matched case-control study, risk profiling by differential epitope binding identified an NPC high-risk group that distinguished 17 of 20 pre-diagnostic sera with an odds ratio of 243.67. However, EBNA1 SeroStrip-HT exhibited a slight sensitivity loss which can be attributed to three NPC sera (0.06, 1.80, and 2.07 years pre-diagnosis) that fell below assay cutoff for the mamEBNA1 dGAr analyte. Successful EBV genotyping of two false-negative samples (1.80 and 2.07 years pre-diagnosis) indicated that both harbored the EBV Akata variant by majority in the blood indicating that the selected analyte sequence was not a limitation in these measurements. Both saliva and plasma have been used for EBV genotyping to reflect the species in the NPC tumor (35,36). The three false-negative samples previously testing positive by the prototype assay represents a limitation of the reduced sensitivity of EBNA1 SeroStrip-HT. One method to improve sensitivity might be to genotype paired mucosal secretions, but nasal aspirates or saliva were not available from the cohort participants (37). Increasing the quantity of analyte may be one method to restore sensitivity without compromising specificity, but the analyte concentration must remain within the linear range of detection.

Only one case fell within the intermediate-risk profile in the samples evaluated within 4 years to diagnosis. A greater number would be identified if time to diagnosis was expanded up to 8 years as previously described (19). Healthy control subjects profiled as high- or intermediate-risk had a median follow-up time of >14 years which indicated that undiagnosed NPC within the control group was unlikely. These profiles provide a practical framework for stratifying screen-positive individuals who lack endoscopic or radiologic evidence of disease, thereby helping to address the major limitation of low PPV in current EBV serologic screens.

IgA to the dGAr analytes were highly correlated between mammalian- and insect-derived proteins (R² = 0.9934 among cases and 0.9848 among controls). This indicates the linear epitopes are preserved regardless of the host cell background. EBNA1 is known to undergo several post-translational modifications (PTMs) during lytic and latent infection (38), yet the comparable performance of the two expression systems suggests that the PTMs are either not a criteria for IgA recognition or are sufficiently conserved in the baculovirus-Sf9 system. Consequently, insect-derived EBNA1 offers a practical route for large-scale production that has precedence for immunologic studies (31,39), but has not been applied to population screening for NPC risk. Similar NPC risk assays often utilize *E. coli*-produced EBNA1 analytes such as truncated proteins and/or peptides in the absence of the GAr (14–16). However, the remaining non-repetitive sequences in EBNA1 are known to contain polymorphic residues that can affect antibody detection and therefore test accuracy (19). This concern has not yet been widely addressed for efficacy in serologic screening. Our EBNA1 analytes are derived from EBV Akata which we have previously shown aligns with nearly all tumor-isolated sequences from endemic and non-endemic regions in China (19), but can differ from other at-risk populations such as in Indonesia with only 4 available whole genome sequences (18). With expanding EBV whole-genome sequencing efforts across geographically and ethnically diverse populations, EBNA1 SeroStrip-HT could be readily adapted to the target population.

EBV serology has been shown to increase early-stage NPC detection and reduce mortality (8,9,11). Although screening for EBV plasma cfDNA is not likely to be cost-effective in the United States even in sub- groups with elevated incidence (40), serology screening programs have been deemed cost-effective in Southern China and in targeted ethnic populations in the United States (32,40). Performance of EBNA1 SeroStrip-HT in NPC low-incidence populations has yet to be evaluated but comparison of alternative models of multiplexed EBV serology assays has shown promising results regardless of endemicity in case-control cohorts in Taiwan and the U.K. (16). Further validation of EBNA1 SeroStrip-HT in diverse cohorts will be an essential step towards defining target populations for implementation, while the inclusion of additional EBV analytes such as VCA p18 and BNLF2b analyzed similarly on a SeroStrip platform could be tested for improved sensitivity and/or discrimination of NPC risk.

## Data Availability

Raw and processed data have been archived in a generalist repository, the Open Science Framework, and are available for download.

https://doi.org/10.17605/OSF.IO/34976

## ACKNOWLEDGEMENTS

This study was supported by the United States National Institutes of Health grant No. R21DE033551 (K. Shair). The Shanghai Cohort Study was supported by NIH grants No. R01CA043092, R01CA144034, and UM1CA182876 (J.-M. Yuan). The Singapore Chinese Health Study was supported by NIH grants No. R01CA080205, R01CA144034, and UM1CA182876 (J.-M. Yuan) and the Singapore Strategic Cohorts Consortium Award grants No. 23-1034-A0001 and P2022-02 (W.-P. Koh). R. Satterwhite was supported by NIH training grant No. T32CA186873. B. Warner was supported by NIH training grant No. T32CA186873 and the Hillman Postdoctoral Fellowship for Innovative Cancer Research made possible by the Henry L. Hillman Foundation. We thank Dr. Troy Messick (Wistar Institute, U.S.A.) for helpful discussions on EBNA1 production and purification.

## SUPPLEMENTARY TABLES AND FIGURES

**Supplementary Table 1.** Baseline demographic and lifestyle characteristics of study participants who developed nasopharyngeal carcinoma (cases) and healthy subjects (controls) from the Singapore Chinse Health Study and Shanghai Cohort Study.

| Characteristic | Singapore Chinese Health Study |  |  | Shanghai Cohort Study |  |  |
| --- | --- | --- | --- | --- | --- | --- |
|  | Cases | Controls | *p*<sup>a</sup> | Cases | Controls | *p*<sup>a</sup> |
| Number of subjects (incident NPC) | 13 | 61 |  | 7 | 35 |  |
| Time to diagnosis (year), median (range) | 1.8 (0.4-3.9) | n/a |  | 1.3 (0.1-3.1) | n/a |  |
| Follow up (year), median (range) | n/a | 12.9 (5.5-19.8) |  | n/a | 28.1 (4.4-31.6) |  |
| Age (year), mean ± SD | 58.2 ± 6.6 | 59.2 ± 5.9 | 0.581 | 55.3 ± 3.0 | 55.0 ± 3.2 | 0.822 |
| Body mass index (kg/m<sup>2</sup>) | 22.5 ± 2.4 | 23.3 ± 3.5 | 0.401 | 23.0 ± 5.1 | 22.3 ± 3.5 | 0.654 |
| Highest level of education, % |  |  |  |  |  |  |
| No formal education | 7.7 | 4.9 |  | 0 | 5.7 |  |
| Primary school | 46.2 | 44.3 |  | 14.3 | 31.4 |  |
| Secondary school or above | 46.2 | 50.8 | 0.901 | 85.7 | 62.9 | 0.483 |
| Cigarette smoking, % |  |  |  |  |  |  |
| Never smokers | 53.9 | 57.4 |  | 42.9 | 62.9 |  |
| Former smokers | 0 | 21.3 |  | 0 | 2.8 |  |
| Current smokers | 46.1 | 21.3 | 0.068 | 57.1 | 34.3 | 0.499 |
| Alcohol drinking, % |  |  |  |  |  |  |
| Nondrinkers | 69.2 | 63.9 |  | 57.1 | 62.9 |  |
| Moderate drinkers | 23.1 | 36.1 |  | 28.6 | 34.3 |  |
| Heavy drinkers<sup>b</sup> | 7.7 | 0 | 0.072 | 14.3 | 2.8 | 0.430 |

<sup>a</sup>2-sided *p* values were based on *t* test for continuous variables or chi-square test for categorical variables; <sup>b</sup>Heavy drinking was defined as >3 drinks/day for men and >2 drinks/day for women, and lower levels as moderate drinking

**Supplementary Table 2.** EBNA1 genotype of pre-diagnostic sera that tested negative by EBNA1 SeroStrip-HT.

| Cohort | Years pre-diagnosis | EBNA1 Serotype |  | EBNA1 Genotype |  |
| --- | --- | --- | --- | --- | --- |
|  |  | Prototype Assay<sup>19</sup><br>*mamEBNA1 dGAR*<sup>a</sup> | EBNA1 SeroStrip-HT<br>*mamEBNA1 dGAR*<sup>b</sup> | Total Reads | % Reads by Residue 487 |
| SCS | 0.06 | Positive | Negative | 143,372 | 99% 487V<br>1% 487A |
| SCHS | 1.80 | Positive | Negative | 137,091 | 97% 487V<br>3% 487A |
| SCS | 2.07 | Positive | Negative | ND | ND |

SCS; Shanghai Cohort Study, SCHS; Singapore Chinese Health Study, ND; not detected, 487V or A; EBNA1 residue at position 487 relative to EBV GenBank ref. V01555.2
<sup>a</sup>Purified EBNA1 dGAR from mammalian cells, <sup>b</sup>Whole-cell lysate from mammalian cells containing EBNA1 dGAR

**Supplementary Figure 1.**
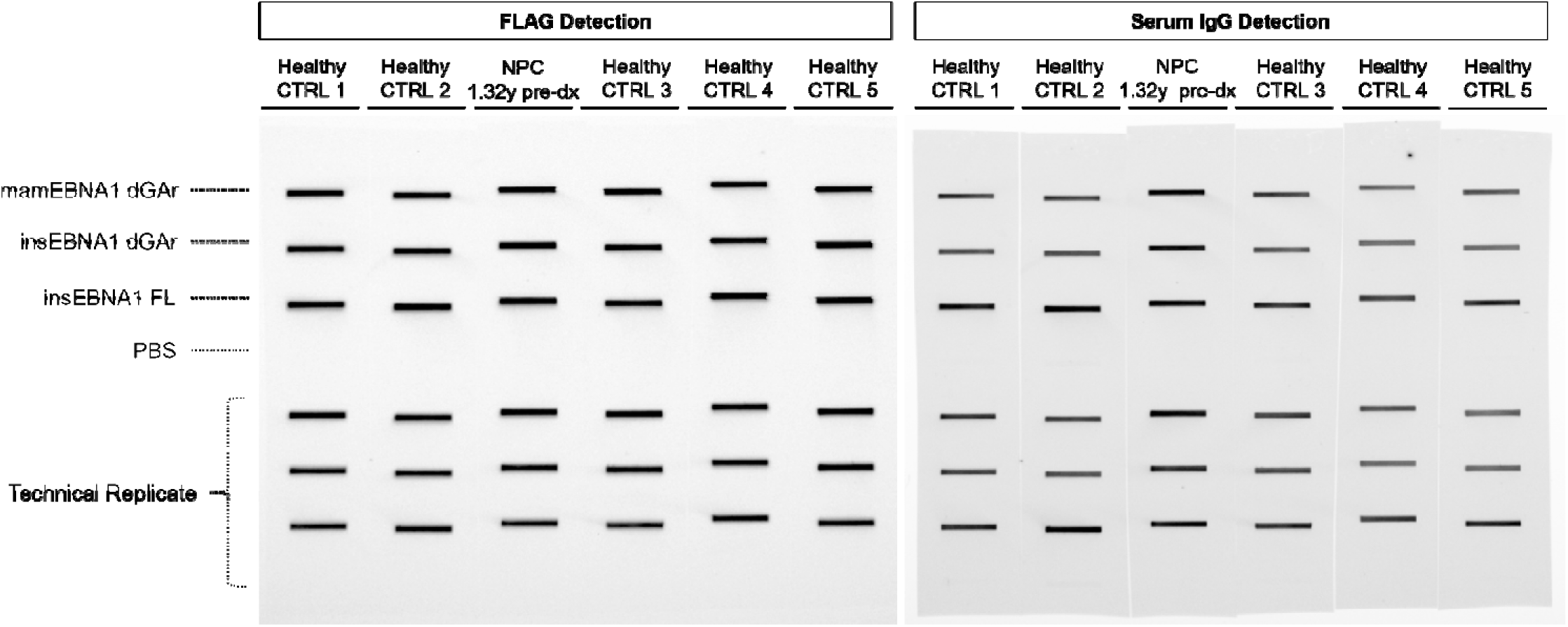
EBNA1 SeroStrip-HT example of FLAG channel participant EBNA1 IgG detection. Protein analytes were loaded in duplicate onto nitrocellulose including EBNA1 deleted for the glycine-alanine repeat produced in mammalian cells (mamEBNA1 dGAr) or insect cells (insEBNA1 dGAr), and full-length EBNA1 produced in insect cells (insEBNA1 FL). Phosphate-buffered saline (PBS) was included as a background control. An N-terminal FLAG-tag permitted fluorescent visualization of each analyte. The IgG channel of the pre-diagnostic (pre-dx) NPC sera 1.32 years to diagnosis is displayed with 5 matched healthy control subjects (see Figure 1 for IgA detection). Densitometry was used to quantify participant antibody detection intensity of each analyte.

**Supplementary Figure 2.**
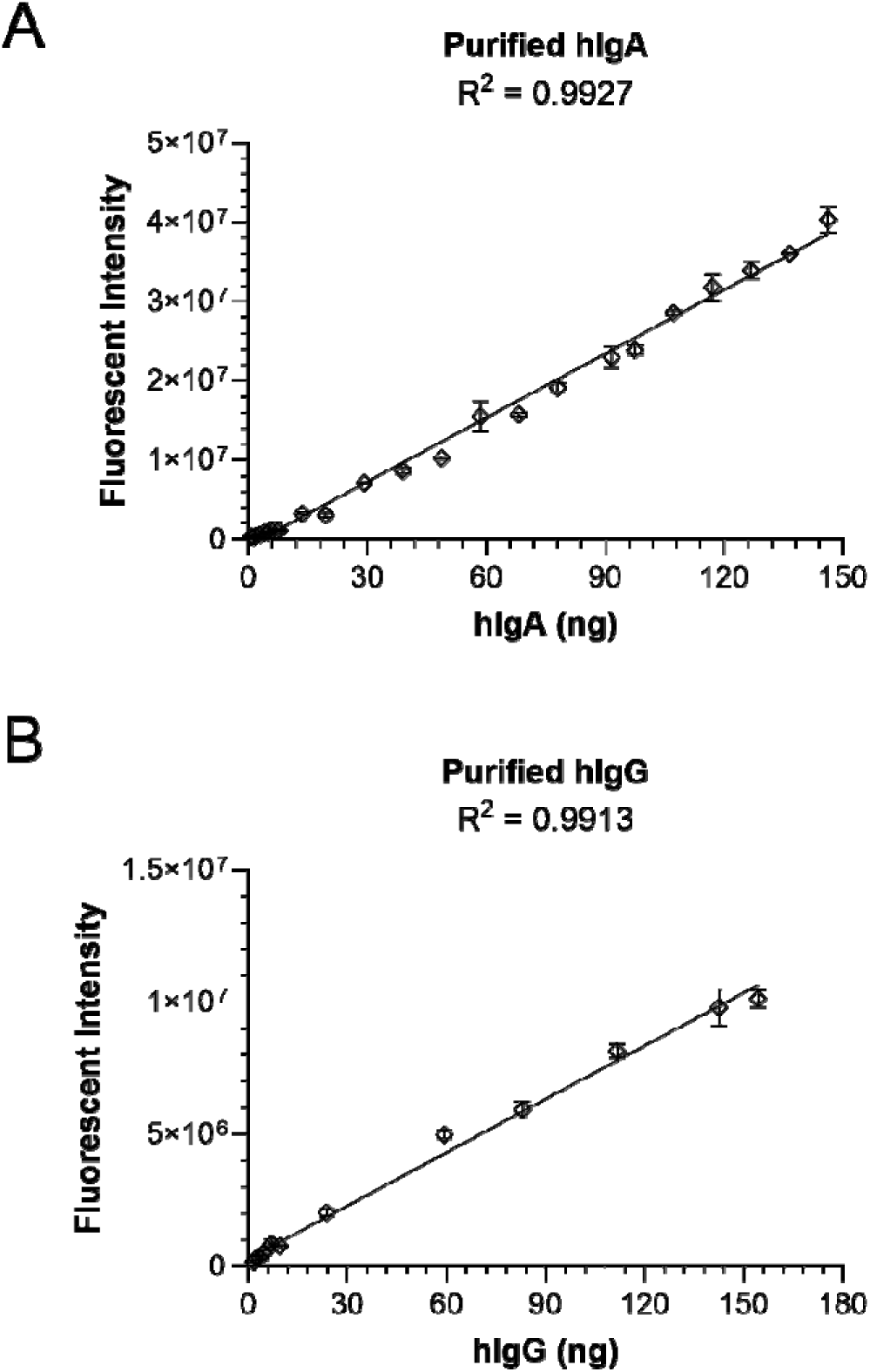
Standard curves for purified human IgA and IgG. **(A)** Known quantities of purified human IgA monomers were loaded onto a nitrocellulose membrane, fluorescently stained, and measured by densitometry to produce a standard curve for absolute quantification of EBNA1 IgA. The same was done for **(B)** purified human IgG monomers to generate a standard curve. R^2^; goodness of fit.

**Supplementary Figure 3.**
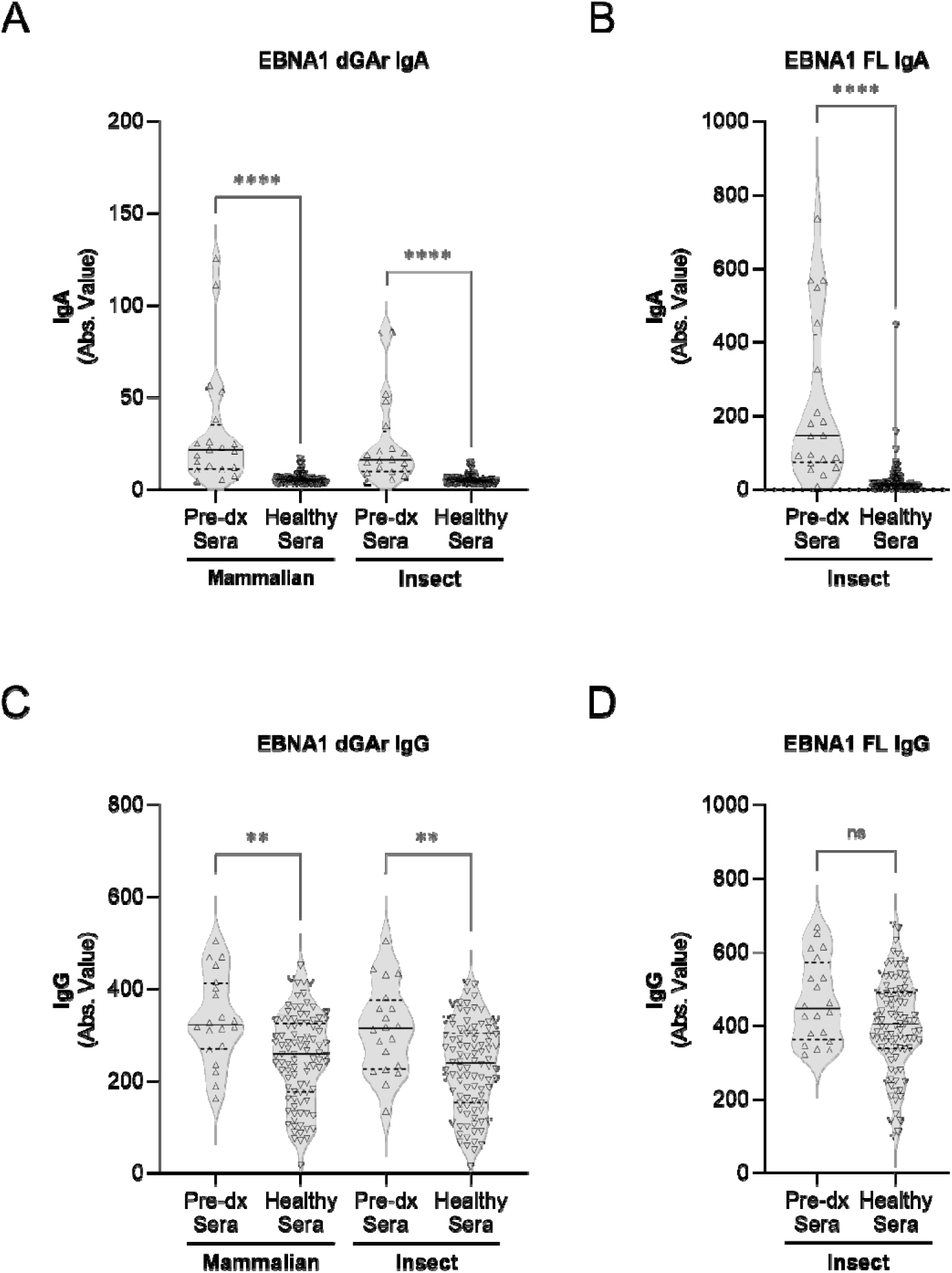
EBNA1 SeroStrip-HT IgA and IgG detection across pre-diagnostic and healthy sera. Pre-diagnostic sera (n=20) and healthy (n=96) sera were measured for **(A)** IgA against EBNA1 deleted for the glycine-alanine repeat produced in mammalian cells (mamEBNA1 dGAr) or insect cells (insEBNA1 dGAr), **(B)** IgA against full-length EBNA1 produced in insect cells (insEBNA1 FL), **(C)** IgG against mamEBNA1 dGAr or insEBNA1 dGAr, **(D)** IgG against insEBNA1 FL. Within violin plot, Solid horizontal bar; median, Dotted horizontal bar; upper and lower quartiles. ****; *P*<0.0001, **; *P* <0.005, ns; not significant.

**Supplementary Figure 4.**
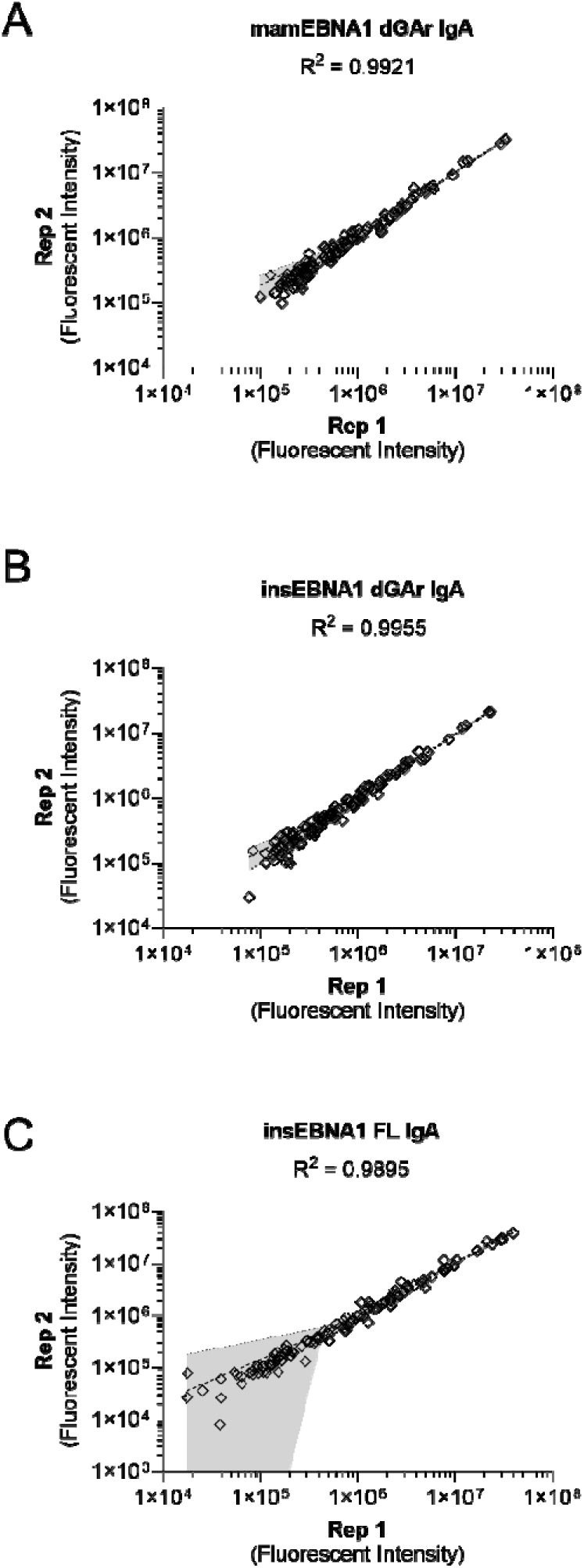
Correlation of Replicate 1 and 2 IgA detection for each EBNA1 analyte. The fluorescent intensity of Replicate 1 (x-axis) compared to Replicate 2 (y-axis) IgA detection of (**A**) mammalian-derived EBNA1 deleted for the glycine-alanine repeat (mamEBNA1 dGAr), (**B**) insect-derived EBNA1 deleted for the glycine- alanine repeat (insEBNA1 dGAr), and (**C**) insect-derived full-length EBNA1 (insEBNA1 FL). R^2^; goodness of fit, Shaded region; 95% confidence band.

**Supplementary Figure 5.**
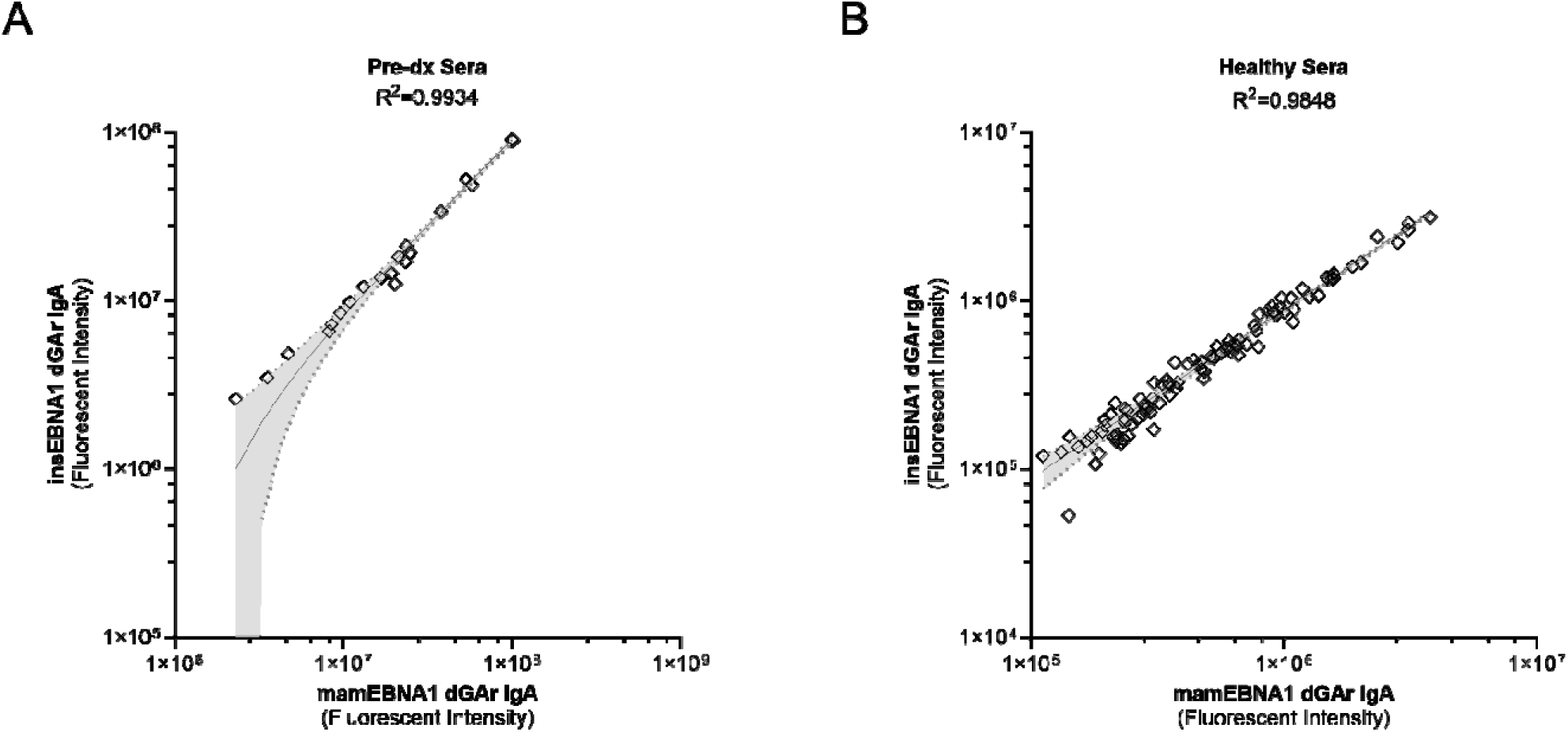
EBNA1 IgA detection correlation between mammalian- and insect-derived proteins. The fluorescent intensity of IgA detection of mammalian-derived EBNA1 dGAr (x-axis) compared to insect-derived EBNA1 dGAr (y-axis) in pre-diagnostic sera (left) and healthy sera (right). R2; goodness of fit, Shaded region; 95% confidence band.

**Supplementary Figure 6.**
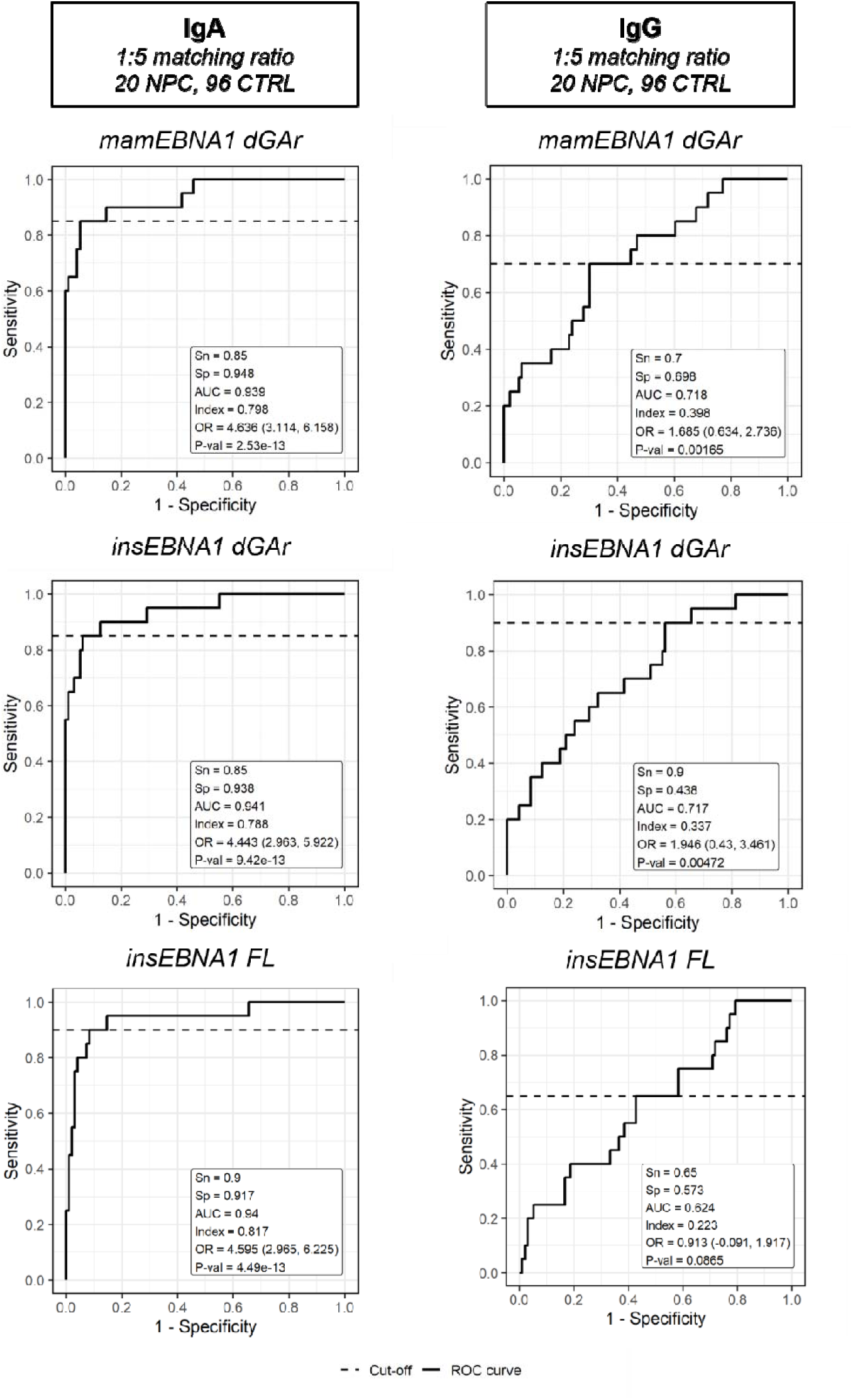
Receiver Operating Characteristic curves for EBNA1 analytes in a 1:5 case-control matching ratio. Receiver operating characteristic curves were generated for IgA (left) and IgG (right) detection of EBNA1 analytes from 20 pre-diagnostic sera and 96 healthy control sera to identify cutoffs for optimal sensitivity and specificity determined by maximal Youden’s index. Cutoff (abs. value), mamEBNA1 dGAr IgA; 10.47, insEBNA1 dGAr IgA; 9.29, insEBNA1 FL; 51.96, mamEBNA1 dGAr IgG; 307.60, insEBNA1 dGAr IgG; 215.60, insEBNA1 FL; 424.30.

**Supplementary Figure 7.**
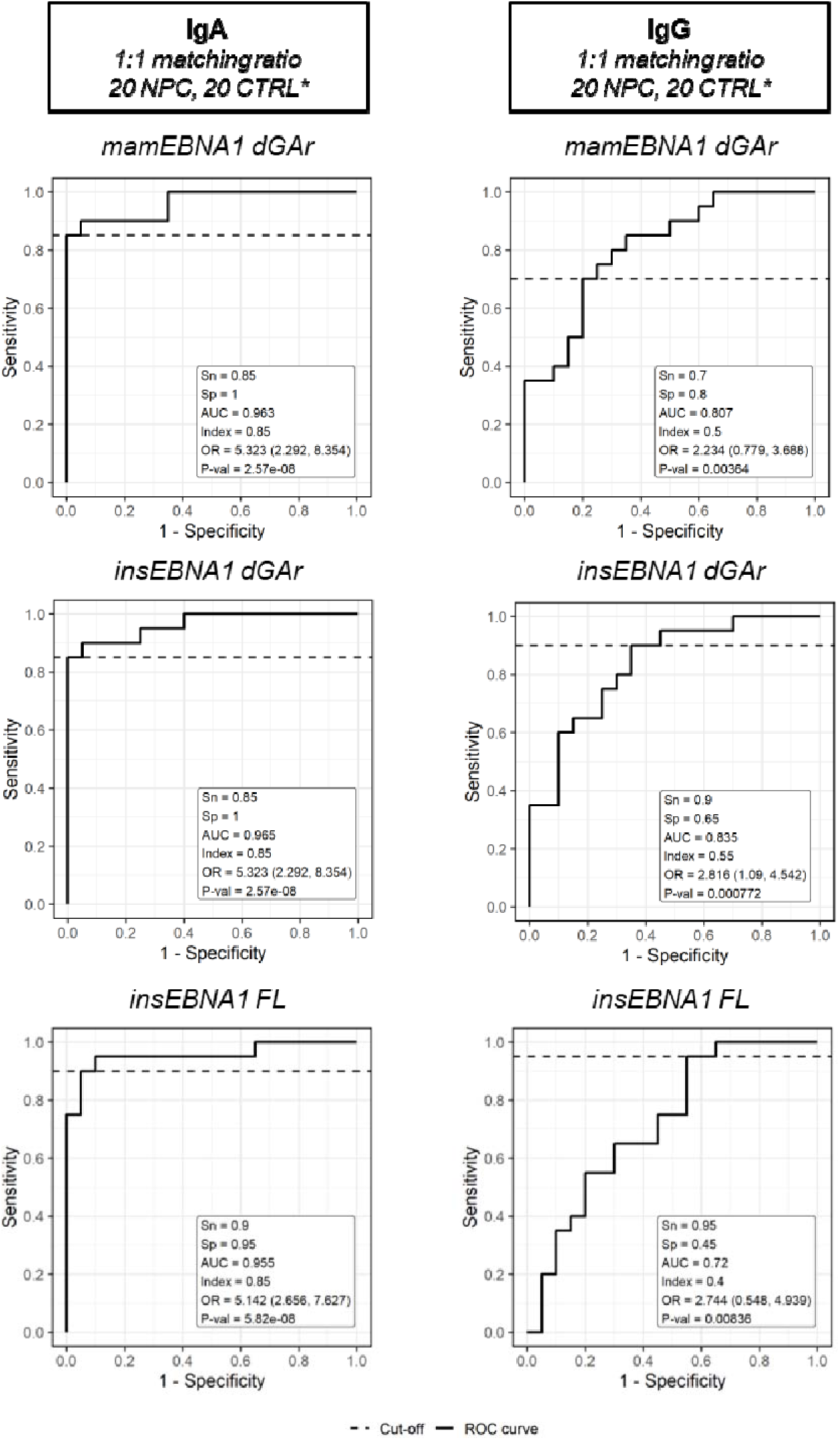
Receiver Operating Characteristic Curves (ROCs) for EBNA1 analytes in a 1:1 case-control matching ratio. Receiver operating characteristic curves were generated for IgA (left) and IgG (right) detection of EBNA1 analytes from 20 pre-diagnostic sera and 96 healthy control sera to identify cutoffs for optimal sensitivity and specificity determined by maximal Youden’s index. Cutoff (abs. value), mamEBNA1 dGAr IgA; 9.60, insEBNA1 dGAr IgA; 8.42, insEBNA1 FL; 50.67, mamEBNA1 dGAr IgG; 307.60, insEBNA1 dGAr IgG; 214.90, insEBNA1 FL; 331.00. *1:1 case-control matching preserved from Warner *et al.* Clin Can Res (2024), ref.19.

**Supplementary Figure 8.**
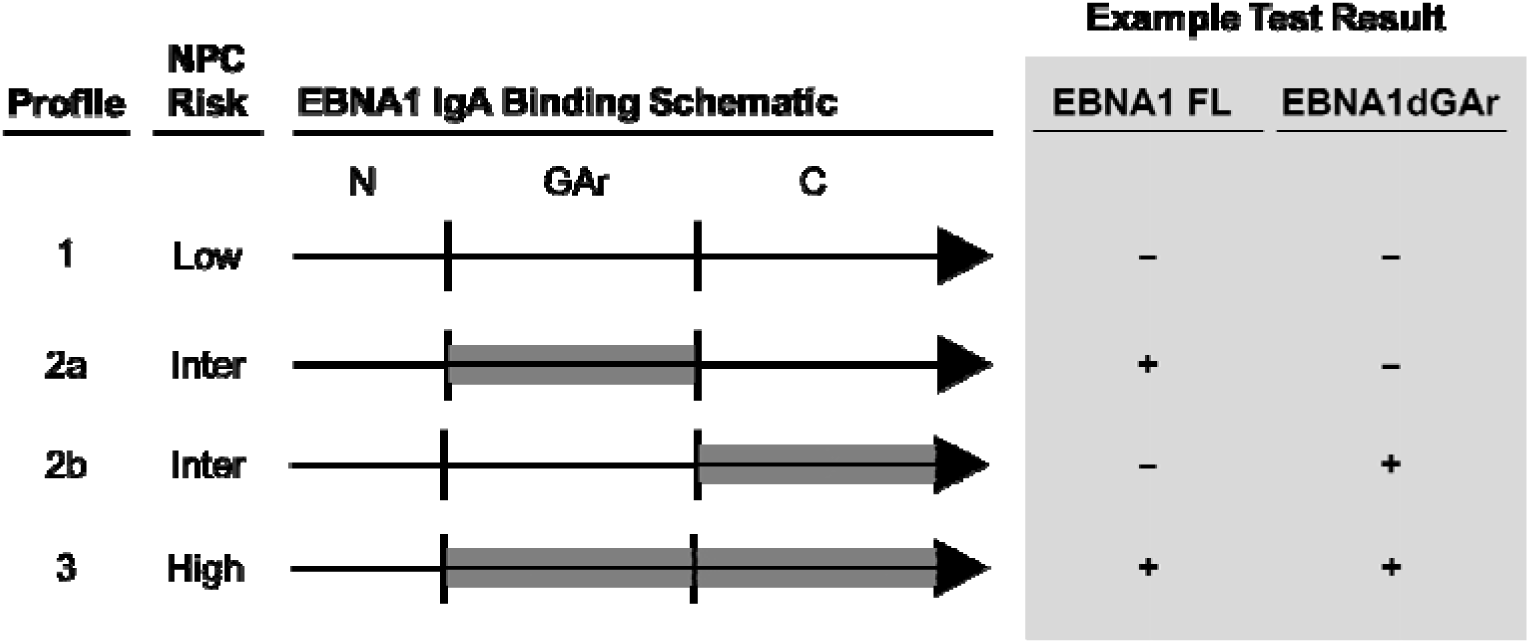
NPC risk profiles based on IgA detection of specific EBNA1 epitopes. Schematic EBNA1 epitope recognition by IgA and risk of NPC diagnosis. Four profiles were established based on IgA against mamEBNA1 dGAr and insEBNA1 FL in a simultaneous testing model. Example test result shown with positive (+) indicating value above cutoff and negative (-) indicating below cutoff. Cutoff (abs. value) from 1:5 matching ratio analysis; mamEBNA1 dGAr; 10.47, insEBNA1 FL; 51.96.

